# Efficient patient-level health economic modelling in Excel without VBA: A Tutorial

**DOI:** 10.1101/2025.06.18.25329835

**Authors:** Rob Blissett, Will Sullivan, Inola Subban, Adam Igloi-Nagy

**Affiliations:** Maple Health Group, LLC, New York, NY, United States of America

## Abstract

Cohort-level models in Microsoft Excel® remain the standard for cost-effectiveness modelling to inform health technology assessment (HTA), despite calls and rationale for more flexible approaches. Their limited ability to capture patient-level characteristics can, in the presence of patient heterogeneity or the need to track patient characteristics to accurately capture a technology’s implications, introduce bias. Their continued prevalence is explained by key stakeholders’ familiarity with spreadsheet software, and the lower computational burden of cohort-level versus patient-level models. However, contemporary Excel functions have opened up possibilities for efficient calculations within native Excel that enable more flexible, patient-level approaches to be implemented in familiar spreadsheet-based software. Therefore, this tutorial aims to provide step-by-step guidance on how to implement a previously published and freely available individual-level discrete event simulation (DES) in Excel, using contemporary Excel functions and without any Visual Basic for Applications (VBA) code.

**Key Points for Decision-Makers:**

- Perceived and real requirements for cost-effectiveness models for HTA to be built in Excel may have led to overuse of cohort-level approaches, with probable bias implications for HTA decision-making.
- Contemporary Excel functions now allow the efficient implementation and execution of patient-level model calculations within native Excel, without any VBA code. Such capabilities may reduce technical barriers across key stakeholders, enhance transparency, and ultimately lead to improvements in HTA decision-making.
- This tutorial demonstrates provides step-by-step guidance on how to implement an efficient patient-level cost-effectiveness model in Excel without any VBA, with an executable model example included as supplementary material.

## 1. Introduction

Cost-effectiveness models for health technology assessment continue to typically comprise cohort-level analyses in Microsoft Excel, despite calls for use of more flexible models and software [1]. Key advantages of more flexible, patient-level models include the ability to readily track patient history and capture patient heterogeneity. If either of these factors are important for accurately modelling the effects of treatment upon outcomes of interest (e.g., quality adjusted life-years (QALYs), healthcare costs), failure to accurately account for them will lead to biased results and potentially decision error risk in health technology appraisal (HTA)[2]. Accounting for patient history and heterogeneity within cohort-level models is possible, for example through tunnel states and subgrouping, but cumbersome. Often, simplicity is traded for accuracy.

Why would a trade-off for simplicity over accuracy be routinely accepted by stakeholders in HTA processes, such as that of the National Institute for Health and Care Excellence (NICE), when the consequences of decision error are counted in population health outcomes? The answer may lie in the near-ubiquitous nature and historical limitations of Microsoft Excel®. Within and outside of HTA, logic and calculations in Excel spreadsheets are considered accessible and transparent, relative to any code-based language. This has led to key skills in building, critiquing and accessing health economic models being developed in Excel spreadsheet logic at the cost of broader development of such skills in code-based, more flexible software options such as R. Even use of Excel’s underlying scripting language (Visual Basic for Applications®; VBA) is typically restricted to programming repeated process in HTA modelling, for example in automating sensitivity analyses. Further, key stakeholders – namely, the pharmaceutical companies commissioning cost-effectiveness models for HTA applications – sometimes mandate their vendors or internal teams to build cost-effectiveness analyses in Excel, owing to internal company rules permitting use of only a limited range of subscribed, closed-source software. In short, Excel remains the software of choice for HTA modelling and accessible modelling in Excel has until now, broadly, meant cohort-level modelling.

An attempt to broaden the typical range of HTA models beyond cohort-level types was made previously, through what was described as discretely integrated condition event (DICE) methodology, but this approach suffered from the same key limitation as previous attempts to build patient-level discrete-event simulations (DESs) or discrete-time simulations (microsimulations) within standard spreadsheet software: slow execution time [3]. Application of DICE methodology in a 2017 National Institute for Health and Care Excellence (NICE) technology appraisal led to run-time criticism from the Evidence Review Group and replication of the model in VBA code to improve run speed mid-appraisal [4]. The slow run time of such models is explained by the need for VBA interaction with Excel spreadsheets at each calculation point; this becomes impractical when the requirement is to generate stable mean probabilistic results for a sufficient simulated sample applied to even a relatively simple patient-level HTA model. However, some more recent developments in Excel functionality may have opened the door to efficient and transparent patient-level HTA models in Excel spreadsheets.

Excel 365 includes previously unavailable dynamic array functions that we have found to be well-suited for efficient specification of patient-level HTA models without any VBA. Of note, the LAMBDA function, introduced in 2021, allows users to define custom functions within Excel cells, without reliance on VBA or external scripting support [5]. Use of LAMBDA in combination with another Excel 365 function, REDUCE, enables efficient, simultaneous calculation of patient-level outcomes, as we illustrate in this tutorial [6].

Given the relative recency of these new functionalities, their application in constructing economic models have been limited. While the general aspects of applying new Excel functions in health economic models have been disseminated, we are unaware of any previous applications of such functionality in models informing HTA submissions [7]. A targeted search was performed for prior examples of economic models leveraging the LAMBDA function; we executed a search of MEDLINE (via PubMed) using search queries to identify any type of economic model that used the LAMBDA function in Excel, with no limits applied to language or time of publication. Combining “lambda” and “Excel” as search terms, there was a single identified hit, which proved to have no relevant connection to our methods [8].

This tutorial aims to provide step-by-step guidance on implementing an efficient, VBA-free patient-level model in Excel, using a reference case DES as a case study [3]. It is designed for those who are familiar with health economic models built in Excel. The remainder of this paper comprises three parts. The first describes the design of the model, with step-by-step instructions on implementation. The second compares key results from the replication exercise, in terms of run-time and accuracy. The third discusses advantages and limitations of the approach in HTA applications, considering potential implications and future directions of research.

## 2. Model Structure and Technical Implementation

This section outlines the structure and implementation of DES model developed in Microsoft Excel to evaluate the long-term clinical and economic consequences of hip fractures. The model tracks patient trajectories over time, capturing the occurrence and impact of fracture-related events. Using modular logic and Excel’s Advanced Formula Environment (AFE), which is part of the Excel Labs software add-in, the simulation supports transparency, reproducibility and full traceability of patient-level outcomes [9]. The following subsections describe the model’s conceptual framework, software architecture and simulation mechanics.

### 2.1 Conceptual Framework and Simulation Logic

The hip fracture model is structured according to the framework described in NICE DSU Technical Support Document 15 [2]. It is designed to capture the clinical and economic consequences of skeletal fragility over time. Individuals enter the model without prior fracture and face risks of vertebral fracture (up to two occurrences), hip fracture, and all-cause mortality. Each event is associated with a decrement in health-related quality of life (HRQoL) and an increase in healthcare costs. Additionally, hip fractures cause an immediate increase in the risk of death. Events occur at stochastically determined times, and the model advances by processing the earliest predicted event, updating the simulated individual’s health status, and accruing costs and QALYs over each interval. The simulation continues until death or the end of the specified time horizon, enabling the evaluation of interventions that modify fracture risk or outcomes. Each individual is simulated sequentially and then results are aggregated across the cohort to generate the results of interest.

### 2.2 Overview of the Model Implementation Approach

An Excel-based DES model was developed using Excel’s AFE, which enables the creation of named, modular functions using the LAMBDA syntax [5]. These functions are stored via the

Name Manager, allowing structured model logic to be composed using native Excel constructs such as LET, HSTACK, VSTACK, and INDEX. We provide an overview of the key functional programming components, particularly LAMBDA and REDUCE, in Section 2 of Online resource 1 [6]. The model file is available as part of the online supplementary materials.

Using these tools, the need for VBA is eliminated while transparency and auditability is preserved [10]. In the example DES framework, each individual is represented by a vector of state variables including accrued costs and QALYs, time of the last event, and scheduled times for future events. The simulation advances iteratively by identifying the next chronological event, applying its effects, and updating the individual’s state. This is implemented using REDUCE over a generated sequence, with each step invoking a transition function that processes a single event and appends the resulting state to the simulation history using VSTACK.

A death flag embedded within the state vector allows early termination of the loop when death occurs. This structure ensures modularity across event types, consistent handling of time-dependent outcomes, and full traceability of the simulated patient history. The next section provides a step-by-step explanation of how the model is implemented within Excel using these components, along with code snippets and references to the full function architecture.

### 2.3 Implementation Steps and Event Handling

In the current model, each row of the simulated cohort corresponds to a single patient, with five random numbers per individual controlling the stochastic elements of their pathway: time to first vertebral fracture, hip fracture, second vertebral fracture, all-cause death, and a hip fracture-related mortality draw. We developed a simulation driver function, runSim (see **Table 1** of Online resource 1, row 1) which is applied on a row-by-row basis, with each row’s random draws and global parameters passed as inputs (Fig. 1).

**Table 1.** Results from three implementations of the hip fracture model (10,000 patients)

|  | Current |  | TSD 15 VBA |  | TSD 15 DICE |  |
| --- | --- | --- | --- | --- | --- | --- |
|  | Comparator | Intervention | Comparator | Intervention | Comparator | Intervention |
| **Undiscounted Results** |  |  |  |  |  |  |
| Cost | £8,970.90 | £8,718.10 | £8,970.90 | £8,718.10 | £8,968.10 | £8,747.59 |
| QALY | 7.375 | 8.140 | 7.375 | 8.140 | 7.329 | 8.100 |
| **Discounted Results** |  |  |  |  |  |  |
| Cost | £6,860.42 | £6,835.11 | £6,860.42 | £6,835.11 | £6,872.82 | £6,858.47 |
| QALY | 6.086 | 6.631 | 6.086 | 6.631 | 6.604 | 6.053 |
| **Model Run Time** |  |  |  |  |  |  |
|  | < 10 seconds |  | 1.5 seconds |  | ~ 25 minutes |  |

While the model’s underlying formulas follow standard Excel syntax, the use of “:=“ in the Boxes below is a notational convention used for readability in this manuscript and is not valid Excel syntax. Additionally, some code snippets include ellipses (…) to indicate omitted arguments or logic that is defined elsewhere. This is used to reduce repetition and maintain focus on the key structure.

These functions are used within runSim to sample times from the Weibull and Normal (**Box 1**) distributions [11].

#### Box 1

**Sampling Helper Functions**

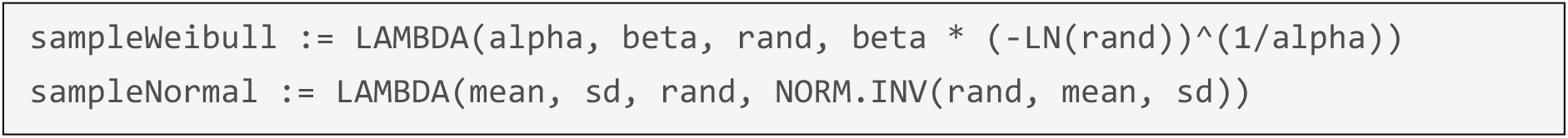

**Box 2** contains the next step of the simulation logic. After sampling, the simulation initializes the individual’s state using a 12-element vector created with HSTACK, capturing time, utility, accrued outcomes, event schedules, and status flags. The model then uses REDUCE to iteratively apply a transition function over a fixed sequence, updating the state at each step. The most recent state is checked for death, and if the individual is still alive, the next event is processed and appended using VSTACK. This structure allows the simulation to advance event-by-event and terminate early upon death, while preserving a full record of the patient’s history.

#### Box 2

**Initializing Patient State and Starting the Simulation Loop**

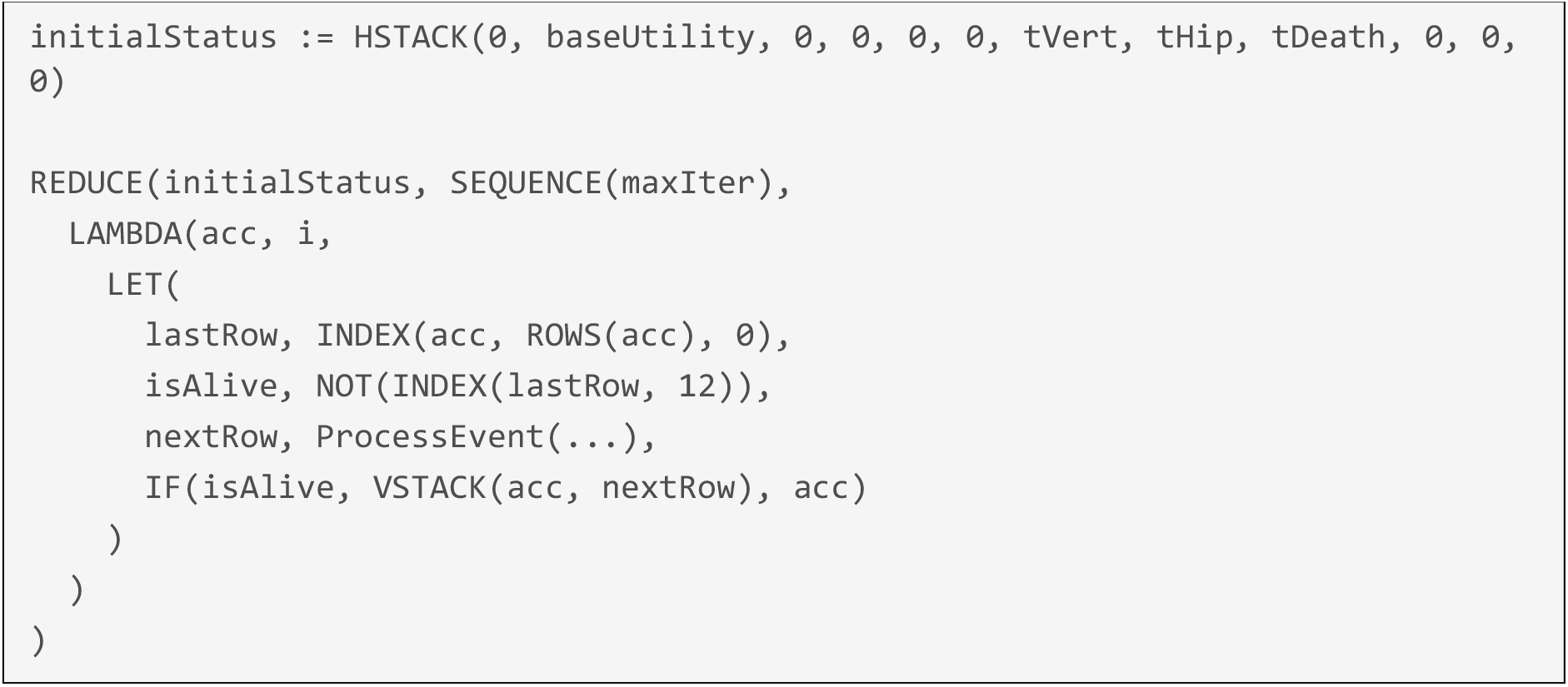

The core event-routing logic is encapsulated in the ProcessEvent function (**Box 3**). This function determines which event occurs next by comparing scheduled event times, and then routes execution to the correct handler function using CHOOSE.

#### Box 3

**Routing to Event Handlers Based on Chronology**

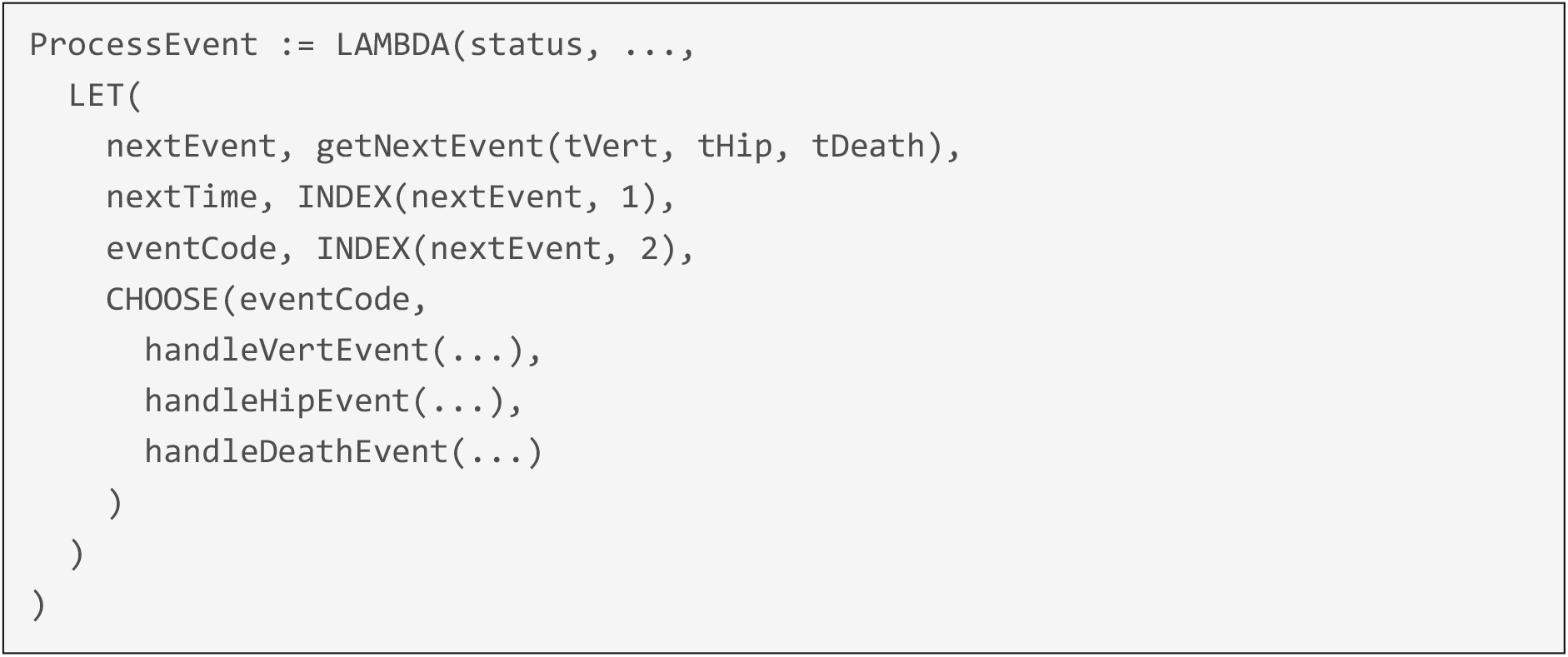

Each event handler is responsible for updating state elements: it accrues QALYs and costs based on time elapsed, applies utility changes, schedules future events where necessary, and updates event flags. For example, in **Box 4**, vertebral fracture affects utility only after the first event, and a second fracture is scheduled if applicable.

#### Box 4

**Event Handler: Vertebral Fracture**

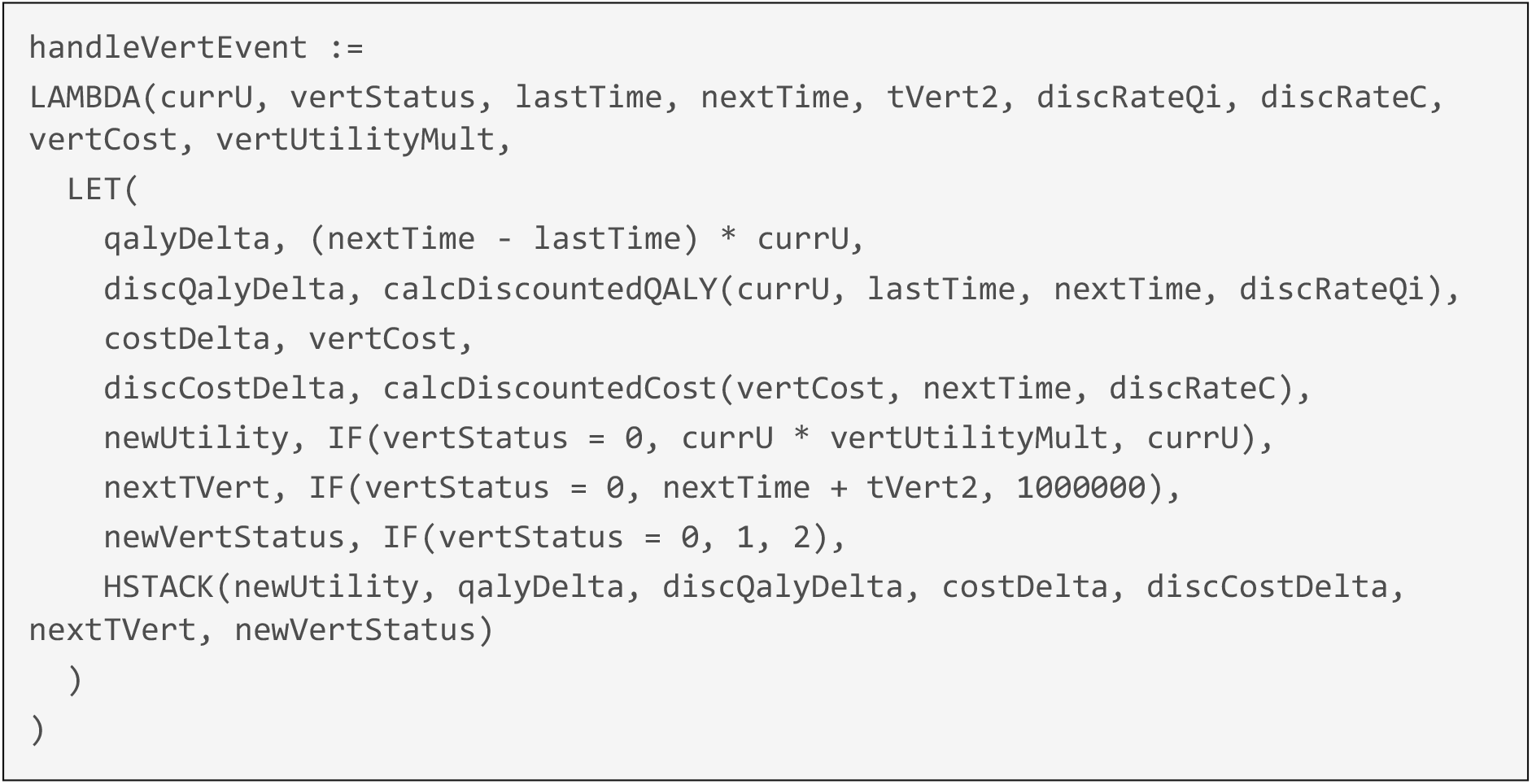

In the last component of runSim logic show in **Box 5**, after all events are processed (or death occurs), the simulation extracts the final patient state using INDEX. This final row includes all necessary outputs for that individual, which are returned in a compact five-element summary vector.

#### Box 5

**Final Output Extraction**

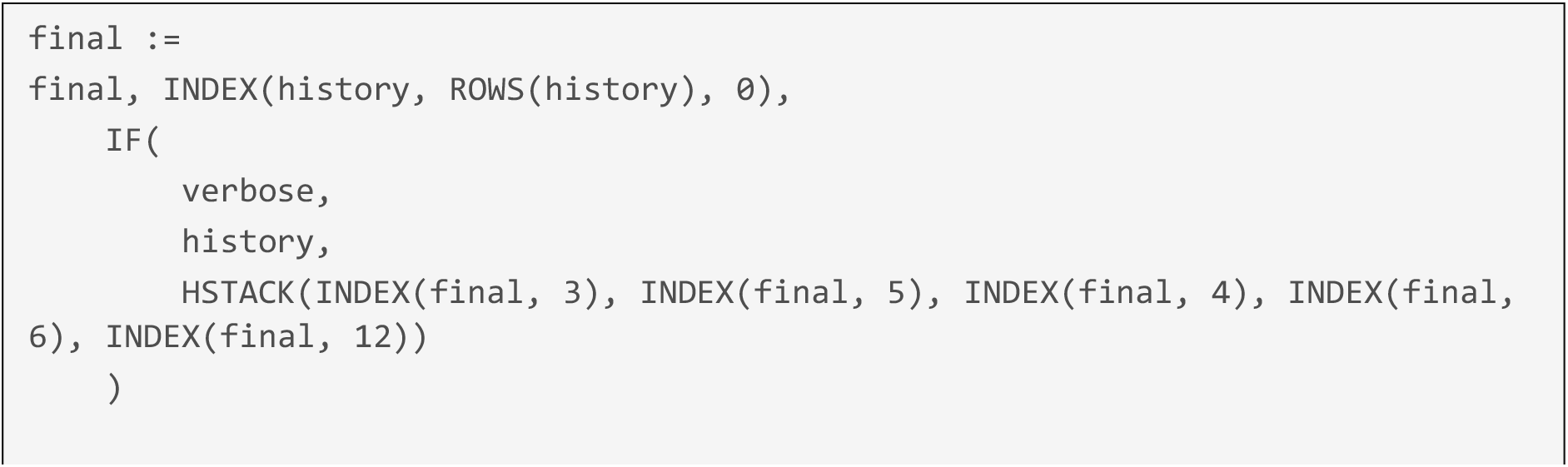

These five-element outputs (QALYs, discounted QALYs, costs, discounted costs, and death flag) are stored across a matrix of patients, with separate arrays for intervention and comparator arms. Aggregation is performed using standard Excel functions (e.g. AVERAGE). Full patient histories can be generated by setting the verbose argument to TRUE (see for example in **Fig. 1**. of Online resource 1. **Table 1** in Online resource 1 provides a full mapping of function inputs, outputs, and dependencies for reference.

**Fig. 1.**
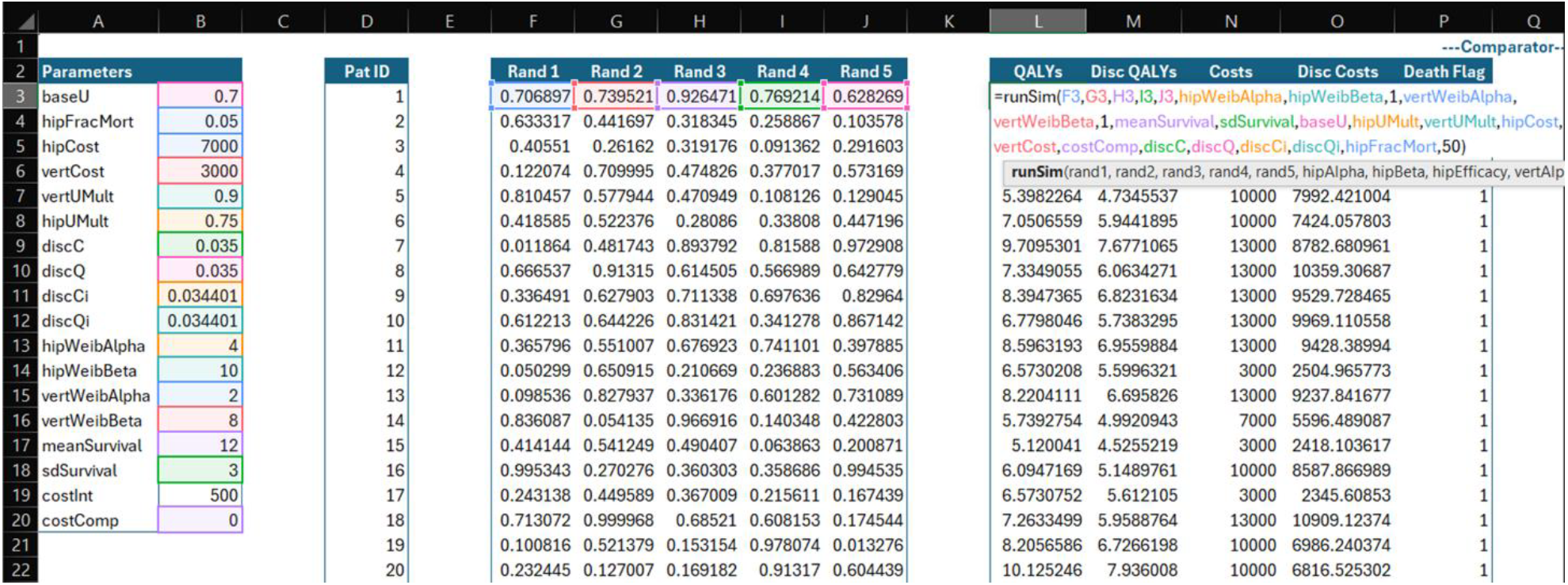
Structure of the Excel-based individual-level simulation.

## 3. Model Replication and Comparative Results

**Table 1** presents a comparison of model outcomes generated using three implementations of the hip fracture simulation: the current Excel-based approach using LAMBDA and REDUCE; the original VBA-based implementation from NICE DSU TSD 15; and version implemented using the DICE methodology also drawn from TSD 15 documentation [2]. Results are reported for 10,000 simulated patients under both comparator and intervention strategies. Across all implementations, cost and QALY outcomes are closely aligned, with only minor differences due to stochastic variation between the DICE model and the other two approaches. In our current model we hard coded identical random number streams to that used by the VBA model, and achieved identical results to the VBA model. Model run time demonstrates a key practical advantage of the LAMBDA-based Excel approach, which generates full-cohort results in under 10 seconds, significantly faster than the 25-minute run time of the DICE implementation, while preserving alignment with the validated TSD 15 outputs.

Following the approach used in TSD 15, we did not generate second-order probabilistic results for probabilistic sensitivity analysis (PSA) here [2]. However, implementing PSA would simply require extending the existing row-by-row simulation to include additional sampling of parameter inputs. The total run time would then scale linearly with the product of the inner and outer PSA loops, reflecting the total number of patients simulated and number of PSA iterations specified.

## 4. Discussion

This tutorial has illustrated how to implement a patient-level cost-effectiveness model within Excel spreadsheet logic, using Excel 365 functions to enable efficient calculations without VBA code. The illustrative example uses a freely available DES previously published in Pharmacoeconomics, allowing the interested reader to compare and interact with the model in R code, VBA code and DICE methodology within Excel, as well as in the efficient no-VBA framework described in this tutorial [3].

The key strength of the approach outlined here over established methods is the removal of meaningful run-time differences between executing a patient-level model in Excel spreadsheet logic and in VBA code. For those practicing health economists with responsibility for building and reviewing cost-effectiveness models for HTA, the approach presented may remove key barriers to broader consideration of more flexible model types. Further, the barriers to learning and teaching cost-effectiveness modelling to students and wider audiences may be reduced, and through transparency gains, quality assurance may improve. Of course, there remain some barriers to learning the techniques and functionality employed in this example, but these are expected to be far lower for the average stakeholder in cost-effectiveness models for HTA, when dealing with native Excel only and not Excel with VBA code. Further, ongoing artificial intelligence developments can democratise such learning, for example through the use of an interactive large language model platform as a modelling assistant.

The ability to implement DES and microsimulation calculations efficiently using Excel functions has the potential to improve the cost-effectiveness analyses informing HTA decisions, which may ultimately reduce HTA decision error. In the modelling of treatments for ophthalmological conditions to inform NICE HTA submissions, for example, there is established precedent for simple cohort-level cost-effectiveness modelling, despite this leading to assumptions with limited clinical plausibility [12]. In one appraisal of a treatment for visual impairment with macular oedema, the submitted analysis was criticised for assuming fixed visual acuity over time in one eye [12]. Patient- and eye-level modelling would more easily allow expected changes to each eye to be captured with fidelity, and the developments outlined in this paper may lower barriers to accurate modelling in future ophthalmological technology submissions to NICE and other HTA bodies.

For more than a decade, there have been repeated calls for cost-effectiveness models to be built in R rather than Excel [13]. Cited benefits have included the ability to run statistical analysis of trial data and model cost-effectiveness within the same environment and the efficiency gains this can bring, particularly with new releases of clinical trial data cuts, as well as the transparency benefits of open-source software. Despite this, Excel has remained the near-universal software of choice for HTA models, due in part to limited industry familiarity with script-based programming languages. The advancements in Excel’s capabilities highlighted by this paper do not overcome every advantage of R for HTA, but they may quieten the debate for some use-cases. For example, execution of a model in Excel remains slower than execution of an equivalent R model (particularly R models leveraging the R package Rcpp, which ports code to C++), but this is only likely to be a practical concern for very complex models. Excel has also been limited in its capacity to allow version control, but this may be changing. Excel Labs’ AFE allows the user to store named functions within modules; these modules can then be stored in ‘gists’ (shareable code snippets which act as separate code repositories) on GitHub, allowing version control and simplifying dissemination [14]. Another widely used argument for R for HTA is that it allows analysis of trial data (e.g., survival analysis for oncology models) to be conducted, stored and updated within the same file as the cost-effectiveness calculations. This is particularly appealing when new, planned pivotal clinical trial read-outs occur.. Given the maturity of packages for this type of survival analysis in R, this remains an advantage. However, modules to perform these tasks could in theory be developed using Excel functions. Excel now also allows the use of Python with spreadsheet formulas, so it is theoretically possible to execute survival analysis within Excel using the Python ‘lifelines’ package.

There are limitations to our case study. As noted, and as per the original NICE DSU TSD 15 models, we have recreated only deterministic results, though we note that this too could be achieved without VBA code and that the implications for probabilistic run times are predictable through simple scaling. While we make our model available as supplementary material for open use, our case study is specific to a previously published DES. The tutorial does not cover other cost-effectiveness model types, though with understanding of the functions and principles described here, applying LAMBDA, REDUCE and other functions we have employed to a microsimulation structure or more typical cohort-level model types should be attainable [15]. As noted above, use of such Excel 365 functionality to capture output of an entire Markov trace within one single-cell formula has been recently disseminated [7].

## 5. Conclusion

This tutorial has provided step-by-step guidance on how to implement a previously published DES in Excel without VBA and with efficient execution speed, using Excel 365 functionality. Wider use of this functionality may broaden the types of cost-effectiveness models used in to address HTA decision problems, with the potential through this and through transparency benefits to improve HTA decision-making.

## Supporting information

Excel Model

## Data Availability

All data in the present study are available in the manuscript and associated supplementary materials

## Notes

**Competing interests:** the authors declare they no conflict of interest relevant to this study.

### Competing Interest Statement

The authors have declared no competing interest.

### Funding Statement

This study did not receive any funding

