## Supplementary File 1 for "Efficient patient-level health economic modelling in Excel without VBA: A Tutorial"

Journal: Pharmacoeconomics

### Online Resource 1.

### 1. Detail of the functions used

*Table. 1. Function dependencies*

| Function Name | Function Purpose | Argument List | Calls or Depends On | Used By / Returns |
|---|---|---|---|---|
| runSim | Main simulation driver that coordinates event sampling, looping, and output extraction | rand1, rand2, rand3, rand4, rand5, hipAlpha, hipBeta, hipEfficacy, vertAlpha, vertBeta, vertEfficacy, meanSurv, sdSurv, baseUtility, hipUtilityMult, vertUtilityMult, hipCost, vertCost, annualCost, discRateC, discRateQ, discRateCi, discRateQi, hipDeathThreshold, maxIter, verbose | sampleWeibull, sampleNormal, REDUCE, ProcessEvent | Outputs QALY, dQALY, cost, dCost, death |
| sampleWeibull | Samples a Weibull-distributed event time | alpha, beta, rand | — | Used within runSim |
| sampleNormal | Samples a Normally-distributed event time | mean, sd, rand | — | Used within runSim |
| ProcessEvent | Determines next event and routes to the appropriate handler | status, tVert2, discRateQi, discRateC, discRateCi, vertCost, hipCost, annualCost, vertUtilityMult, hipUtilityMult, hipDeathRisk, hipDeathThreshold | getNextEvent, handleVertEvent, handleHipEvent, handleDeathEvent | Called by runSim → REDUCE |
| getNextEvent | Identifies the earliest of the three scheduled events | tVert, tHip, tDeath | — | Used within ProcessEvent |
| handleVertEvent | Updates state and outcomes after a vertebral fracture | currU, vertStatus, lastTime, nextTime, tVert2, discRateQi, discRateC, vertCost, vertUtilityMult | calcDiscountedQALY, calcDiscountedCost | Used within ProcessEvent |
| handleHipEvent | Handles utility and cost changes after hip fracture and determines fracture-related death | currU, lastTime, nextTime, discRateQi, discRateC, discRateCi, hipCost, annualCost, hipUtilityMult, hipDeathRisk, hipDeathThreshold | calcDiscountedQALY, calcDiscountedCost, calcDiscAnnCost | Used within ProcessEvent |
| handleDeathEvent | Finalizes state at death and calculates final QALYs and costs | currU, lastTime, nextTime, discRateQi, discRateCi, annualCost | calcDiscountedQALY, calcDiscAnnCost | Used within ProcessEvent |
| calcDiscountedQALY | Computes QALYs over an interval with continuous discounting | u, t1, t2, discRateQi | — | Used in all event handlers |
| calcDiscountedCost | Computes a one-time cost with exponential discounting | cost, t, discRateC | — | Used in vertebral and hip handlers |
| calcDiscAnnCost | Calculates the present value of a stream of annual costs using continuous discounting | cost, t, discRateCi | — | Used in hip and death handlers |

### 2. Overview of LAMBDA/REDUCE

```
=LAMBDA(x, y, x + y)
```

This simple function takes two inputs (x and y) and returns their sum. This can be saved in the spreadsheet using Name Manager, or more easily using the AFE. It then allows a user to call the function in any spreadsheet cell. This is powerful because it allows recursion. Consider the following example. We define a new name, Factorial:

```
=LAMBDA(n, IF(n<=1, 1, n * Factorial(n - 1)))
```

This formula will return 1 when n is 1 but otherwise will return the factorial of the number supplied (e.g., for 5 this is 5 x 4 x 3 x 2 x 1). It should be noted that recursion in a LAMBDA function can only be called up to 254 level deep. In cases where we need to exceed this limit, we can make use of REDUCE (or SCAN or MAP, but these are beyond the scope of this tutorial) for iterative solutions. REDUCE applies a LAMBDA function repeatedly to an array carrying forward an accumulated result. In the factorial example above this becomes:

```
=REDUCE(1, SEQUENCE(5), LAMBDA(a, x, a * x))
```

The SEQUENCE function is a dynamic array function that generates an array of sequential integers, in this case {1;2;3;4;5). The REDUCE function starts with an accumulator (a = 1) and multiplies each x from the sequence into the accumulator.

```
(((1 × 1) × 2) × 3) × 4) × 5 = 120
```

We can save this as a named function for the general case:

```
=LAMBDA(n, REDUCE(1, SEQUENCE(n), LAMBDA(a, x, a * x)))
```

### 3. Patient histories

*Figure 1 – Example of dynamic full patient history output available by calling runSim with verbose set to TRUE*

**Check full patient history**

| Patient look up | |
|---|---|
| Patient ID | 1 |
| Model arm | Comparator |

| Time | Utility | QALY | Cost | Disc QALY | Disc Cost | tVert | tHip | tDeath | vertStatus | hipStatus | deathFlag |
|---|---|---|---|---|---|---|---|---|---|---|---|
| 0 | 0.7 | 0.00 | 0 | 0.00 | 0 | 4 | 8 | 14 | 0 | 0 | 0 |
| 4 | 0.6 | 3.08 | 3000 | 2.85 | 2579 | 7 | 8 | 14 | 1 | 0 | 0 |
| 7 | 0.6 | 4.47 | 6000 | 4.01 | 4969 | 1000000 | 8 | 14 | 2 | 0 | 0 |
| 8 | 0.5 | 5.14 | 13000 | 4.53 | 10345 | 1000000 | 1000000 | 14 | 2 | 1 | 0 |
| 1000000 | 0.5 | 8.23 | 13000 | 6.66 | 10345 | 1000000 | 1000000 | 14 | 2 | 1 | 1 |
